# Error Rates in SARS-CoV-2 Testing Examined with Bayesian Inference

**DOI:** 10.1101/2020.12.17.20248402

**Authors:** P. M. Bentley

## Abstract

A literature review on SARS-CoV-2 reverse-transcription polymerase chain reaction (RT-PCR) is used to construct a *clinical* test confusion matrix. A simple correction method for bulk test results is then demonstrated with examples. The required sensitivity and specificity of a test are explored for societal needs and use cases, before a sequential analysis of common example scenarios is explored. The analysis suggests that many of the people with mild symptoms and positive test results are unlikely to be infected with SARS-CoV-2 in some regions. It is concluded that current and foreseen alternative tests can not be used to “clear” people as being non-infected. Recommendations are given that regional authorities must establish a programme to monitor operational test characteristics before launching large scale testing; and that large scale testing for tracing infection networks in some regions is not viable, but may be possible in a focused way that does not exceed the working capacity of the laboratories staffed by competent experts. RT-PCR tests can not be solely relied upon as the gold standard for SARS-CoV-2 diagnosis at scale, instead clinical assessment supported by a range of expert diagnostic tests should be used.

## 1. Introduction

During the ongoing SARS-CoV-2 pandemic, there have been understandable calls for widely available testing procedures [1]. The primary use cases were:

1. Identifying infected people in the population as early as possible, ideally before symptoms are exhibited, so that measures can be taken to avoid spreading the disease to others.
2. Confirming SARS-CoV-2 infection in patients exhibiting symptoms, so that they can be isolated, treated and/or studied separately from patients with other illnesses.
3. Ruling out SARS-CoV-2 infection, allowing a person to avoid isolation when exhibiting the milder symptoms shared with other infections of the respiratory tract.

A common, moderate cost and efficient SARS-COV-2 test is based around the reverse-transcription polymerase chain reaction (RT-PCR) method. This was, at the time of writing, referred to as the “gold standard” perhaps optimistically. Indeed, efforts to validate serological testing [2] and computerised tomography (CT)-based methods [3] have used RT-PCR as a reference of “confirmed cases” by which to measure other testing methods. RT-PCR is a relatively simple method, requiring a swab sample that is sent to the lab for chemical amplification.

Use case 1 would ideally involve a large number of tests being performed on the general public, and a number of governments have expressed intention to do this at scale. Use cases 2 and 3 are often performed on admission to a clinical facility. Use case 3 is particularly important for critical workers in society, allowing them to return to their duties without fear of spreading the disease [4] and became a deployed strategy in some regions (e.g. the UK) early in the pandemic.

These use cases, and the policies of many governments, assume low error rates from the tests. The reality of any test, unfortunately, is that errors do occur. Moreover, whilst the statistics of testing is a core component of undergraduate scientific education, because even seasoned experts occasionally make statistical mistakes it is worth expending a little patience to cover the ground-work before tackling the main body of the problem.

This article will therefore summarise the known fundamentals of testing and bayesian methods for a general readership. The contributions thereafter are the derivation of a correction method for public data to replace the belief that “a positive test equals a confirmed case”; before a review of clinical literature is combined with these methods. There are logically new implications for SARS-CoV-2 diagnosis; updated probabilities for various prognoses; suggestions for public policy; and the validity of research relying on “confirmed” cases of SARS-CoV-2 that are described as a consequence of this work.

### 1.1. Test Confusion Matrix

A *confusion matrix* conveniently encapsulates the reliability characteristics of a test, shown in table 1. One column holds the positive condition (in this case, “Infected”) and the other column holds the negative (in this case, “Healthy”). Each row corresponds to a test result, either positive or negative. Thus one sees that the confusion matrix is a table of test results that are *true positive (t*_*p*_*), true negative(t*_*n*_*), false positive (f*_*p*_*), and false negative (f*_*n*_). These numbers could be given as tallies of results, or they could be normalised so that each column sums to unity and each matrix element represents a probability of that test result being given for a given infection status.

**Table 1:** Confusion matrix for a generic test.

|  | Infected | Healthy |
| --- | --- | --- |
| Test Negative | $f_n$ | $t_n$ |
| Test Positive | $t_p$ | $f_p$ |

In addition to estimates of variables one must also propagate the uncertainty, error, or statistical accuracy of the values. This usually has the symbol *σ*, and is given by the square root of the counts of the measured quantity. *σ* is widely used in physics, but frequently in medicine one quotes the 95% statistical confidence level, which is ∼ 1.96 × *σ*. Considering a number of positive *n*_*pos*_ and negative *n*_*neg*_ test results from a total of *n* = *n*_*pos*_+*n*_*neg*_ tests, we estimate the probability of returning a positive result, and it’s confidence range, with the following well-known equations:

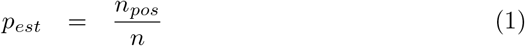

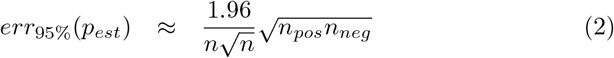

In other words, we have our best estimate of *p*_*est*_, and 95% confidence that *p* actually lies betwen (*p*_*est*_ − *err*) and (*p*_*est*_ + *err*).

The test characteristics are often presented as well-known parameters. The *sensitivity, s*_*e*_, or *true positive rate* (TPR) measures how much of the “infected” column is correctly identified. It is given by:

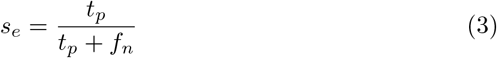

and the *specificity, s*_*p*_, or *true negative rate* (TNR) measures how much of the “healthy” column is correctly identified. It is given by:

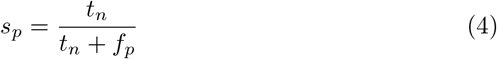

These are related to the *false negative rate* (FNR) and *false positive rate* (FPR) by

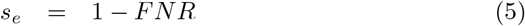

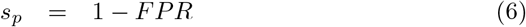

The false positive and false negative rates are to some degree tuneable by the test designer. This can be visualised as a “gain” control on an amplifier. Turning up the gain makes it more likely to catch fainter, positive signals, (false negatives decrease). The “gain” here in the amplification process is therefore correlated very strongly with the statistical sensitivity. However, increasing sensitivity therefore increases the noise (false positive rate increases). Conversely, turning down the gain reduces the noise (false positive rate goes down) but makes it more likely that you miss weaker signals of interest (false negative rate increases).

Test designers therefore try to balance these two effects to minimise risk. ROC curve analysis [5] can be used to tune test procedures quite accurately for a given prevalence. Including cost/benefit analysis in the test design [6] allows one to adjust the sensitivity of the test relative to the disease prevalence, which was summarised very well by Kaivanto [7]. As a side note, it seems that some batches of false positive results are likely to be related to incorrect sensitivity for a particular use case, and not simply statistical anomalies or quality issues.

The confusion matrix in table 1 allows us to write two simultaneous equations for the situation where a number of tests are used in the field. Let us imagine that in a testing programme, *n*_*pos*_ of these tests return positive results, and *n*_*neg*_ return negative results. How many are actually infected? Let us further imagine that, before launching the mass testing programme, one took the essential step of fully mapping the confusion matrix with a thorough clinical study (currently lacking for SARS-CoV-2 testing). One can then establish, from the test result totals, the actual number of infected patients *N*_*i*_. We must first eliminate the number of non-infected or healthy patients *N*_*h*_ from

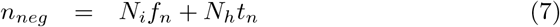

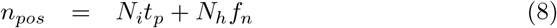

These are simply a symbolic representation of table 1. Solving these simultaneous equations for *N*_*i*_ then yields a simple equation to estimate for the number of patients actually infected with SARS-CoV-2:

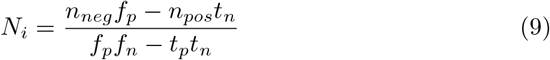

Instead of erroneously reporting *n*_*pos*_ as the number of infected people, *N*_*i*_ gives the accurate result if the test has been properly characterised. This equation is applied in sections 3.1 and 3.2.

### 1.2. Bayes’ Theorem and Base Rate Fallacies

If one would like to use a test to diagnose a patient, or to rule out possible infection so that they can be safely released back into society or a work function, the confusion matrix alone is insufficient. One must also consider the base rate, or prevalence, in the context of the test. For example, a test that has a 90% sensitivity incorrectly clears 10% of those infected. If we imagine an enclosed group, for example a jail, filled with sick patients in their beds, it is intuitive that any test results coming back negative from symptomatic patients in that group should be treated with caution.

Conversely, if one used a test that has a 90% specificity, it still returns a false positive 10% of the time. If one then attempts to screen millions of citizens in an attempt to find individuals with a disease afflicting one in a thousand people, then one intuitively knows that the infected cases will be buried amongst hundreds of thousands of false positive results.

Ignoring the prevalence of the phenomenon for which one is testing is a well known statistical error called the *base rate fallacy*. Taking into account the base rate, and the confusion matrix, one can introduce combinations of probabilities to study common scenarios. For example, whether or not a person has symptoms, and is tested, what is the probability that the person is actually infected, considering that there exist alternative diagnoses with similar symptoms, and that some patients remain symptom free?

The key to tackling these scenarios rapidly, objectively, and conclusively, is Bayes’ theorem. Bayesian inference has been applied in two forms: both using continuous distribution functions or discrete variables. This article uses the latter, i.e. probability functions of Boolean variables of disease evidence *e*. The evidence *e* = 1 could be a positive test result, or exhibition of symptoms, whilst *e* = 0 is the absence of this evidence. The disease status *d* = 1 indicates infection, and *d* = 0⇒ ¬ *d* = 1 indicates the lack of infection. In these terms, Bayes’ theorem is:

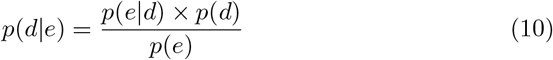

*p*(*d* | *e*) is the conditional probability that we are trying to establish: given the evidence *e*, what is the probability that the person has the disease status *d*? In maths and physics, this is known as the “posterior”, and in the medical community it is known as the “posttest” probability.

*p*(*e* | *d*) is the likelihood of obtaining evidence *e*, assuming that the patient has the disease. If the evidence is a positive test result, and one took all the infected patients who had the disease, then *p*(*e* = 1 | *d* = 1) is the fraction of those patients that would be expected to return a positive test result: it is the true positive rate of the test *t*_*p*_ from section 1.1. If evidence *e* is a symptom of the disease, then *p*(*e* | *d* = 1) is the fraction of infected patients who exhibit that symptom, based on expert clinical studies of the disease.

*p*(*d*) is the (“prior”) probability, or base rate, of any individual having disease status *d*, irrespective of the evidence *e*. In the medical community, it is called the “pretest” probability.

Lastly, *p*(*e*) is the (“marginal”) likelihood of obtaining evidence *e* considering both that the patient may have the disease or may not.

The marginal term *p*(*e*) is conveniently expanded using the law of total probability:

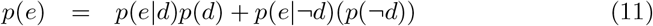

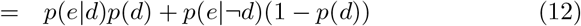

where, for example, *p*(*e* = 1|*d* = 1) is the probability of an infected person yielding a true positive result; and *p*(*e* = 1|¬*d* = 1) is the probability of obtaining a false-positive test result *e* = 1 from a non-infected patient (*f*_*p*_ in section 1.1): these variables can be obtained from tables of test results from clinical studies, as will be shown in tables 2 and 3. Equation 11 demonstrates the method of logical combinations of probabilities. If we imagine events *P* and *Q* that occur independently, with probabilities *p*(*P*) and *p*(*Q*) respectively, then:

**Table 2:** Confusion matrix for the RT-PCR test using data from hospital-administered tests of more than 1000 patients, reported by Ai *et al* [3]. The lower upper two rows show Ai’s reported data, the lower two rows convert these into rates. The ranges in parenthesis correspond to the 95% confidence intervals.

| Test Result | Infected | Healthy |
| --- | --- | --- |
| Negative | 308 | 105 |
| Positive | 580 | 21 |
| Negative | 0.35 (0.32–0.38) | 0.83 (0.77–0.90) |
| Positive | 0.65 (0.62–0.68) | 0.17 (0.10–0.23) |

**Table 3:** Working confusion matrix for the rest of this study. Specificity of 83% will be called “pessimistic”, and 97.4% will be called “optimistic”. These data, obtained from the most recent clinical results in the literature, are used as the *p*(*e* | *d*) and *p*(*e* |¬ *d*) parameters in equations 10 and 11.

| Test Result | Infected | Healthy |
| --- | --- | --- |
| Negative | 0.35 | 0.83 – 0.974 |
| Positive | 0.65 | 0.17 – 0.026 |

- **AND**: *p*(*P ∧ Q*) = *p*(*P*) × *p*(*Q*)
- **OR** : *p*(*P ∨ Q*) = *p*(*P*) + *p*(*Q*)
- **NOT**: *p*(¬*P*) = (1 − *p*(*P*))

One can immediately see, then, why an impressive-sounding test likelihood *p*(*e* | *d*) leads people into the base rate fallacy, *i.e*. forgetting to normalise by multiplying with the base rate *p*(*d*) and dividing by the marginal term *p*(*e*). It is also the current situation facing many with RT-PCR test results, compounded by the use of laboratory rates of sensitivity and specificity rather than those in the field.

The marginal term has one final noteworthy utility, that is to remove the effect of time bracketing of illnesses, symptoms, or statistics gathering. Some rates are given per day, per week, or per year, and the marginal allows us to compare fairly disparate definitions of rates.

Bayes’ theorem can be applied sequentially to multiple scenarios, where the “output” posterior probability of one assessment *p*(*d*| *e*) is used as the “input” prior probability *p*(*d*) for a subsequent test, because combining multiple scenarios with logical **AND** is simply multiplication. This is known as bayesian inference or bayesian updating, where each step adds a new fact that is used to quantiatively adjust the confidence level of the hypothesis. This is also known as Bayesian belief updating — or evidence accumulation — where “today’s posteriors are used as tomorrow’s priors”.

### 1.3. Testing and Policy

Despite being refuted by clinical expert input [8], at the time of writing the strategy of seeking a single negative RT-PCR test result to indicate an absence of infection was in use in some areas. In Sweden, for example, the public health agency — Folkhälsomyndigheten — stated [9] that “Testing people with symptoms of covid-19 who work in socially important activities to be able to rule out disease is important.” Which it is. The organisation then provided links, via another organisation, identifying which jobs fell into this category. It was then up to the regional powers to implement guidelines. At the time of writing, people did not have to isolate after a negative test result once symptoms disappear or after waiting for 7 days [10]. This strategy is a mistake because it ignores false negatives: patients who *are infected* with SARS-CoV-2 but for whom the test result is incorrect.

Meanwhile, the advice from the United States Centers for Disease Control and Prevention stated [11] for a significant part of 2020 that a “positive test result means you have an infection”. The published threshold for detection at 95% confidence by one major supplier of SARS-CoV-2 test kits was 136 copies/mL [12], which evidently leads to confidence in the test results, and by which clinical guidelines have been written that assert laboratory test specifications as being representative of operational specifications [13]. These both assumed, perhaps prematurely, a negligible operational rate of false positives: patients who *were healthy* and for whom the test result was incorrect.

During the writing of this article, the CDC have correctly updated their guidelines [14]. Whilst they still stated that a positive test result “indicates that RNA from SARS-CoV-2 was detected, and therefore the patient is infected with the virus and presumed to be contagious” there were disclaimer clauses encouraging clinical observations and context for positive test results, and that negative test results do not rule out SARS-CoV-2.

The UK guidance, from the country’s National Health Service, specified [15] that a person testing negative did not need to self isolate if “everyone you live with who has symptoms tests negative”, amongst other criteria. However, with a false negative rate of 35%, which is representative, just over 1 in 10 infected households would return negative results for a couple, and more than 1 in 100 would return all negative for a family of four. The UK advice specified further mitigating measures, including that a person who feels sick should still isolate at home, but it did not offer guidelines as to how long.

In contrast, the French labour ministry specified a fairly rigorous quarantine protocol [16]: that anyone who had encountered an elevated risk situation should isolate for 7 days, then take a test. A positive test result required 7 further days of isolation. Even with a negative result, a person who had symptoms must continue isolation until 48 hours after the fever subsides. The public health agency stated that in the case of a negative test the patient should inform the doctor and respect their advice [17]. This is a sensible improvement over the Swedish policy, leaving the possibility open for expert input to rule out false negatives, but it carries possible inconsistency over a range of interpretations and diagnoses.

All of the above is not to say that any specific country, or organisation, is wrong to deploy tests with significant error rates. In an emergency situation, it is correct that any test is better than no test at all. However, SARS-CoV-2 is no longer a new disease: the pandemic has been running for more than a year. It is essential that the error rates are properly understood, to minimise the impact of incorrect test results.

## 2. Existing Literature

It was identified at the early stages of the pandemic that RT-PCR tests used *outside the laboratory setting* were underwhelming when used as a reference for other clinical testing options [3]. Using Ai’s data, one can construct a confusion matrix for RT-PCR tests relative to chest x-ray combined with diagnosis from a qualified medical expert, which is summarised (as an example) in table 2.

One should note that those RT-PCR tests were performed in a clinical setting by a trained medical worker, supported by an expert laboratory. Collecting the test sample is not painful, but uncomfortable because it triggers a gag reflex and a strong negative reaction to an object pushed into the nasal cavity. For a detailed overview and discussion, see Syal [18]. The end result is that for home testing kits, drive-thru facilities, or similar, where the patient or a family member collects the samples, and the processing of the kits is done at scale by non-expert technicians, one should therefore anticipate additional adverse effects — particularly an increase in false negatives.

There has been an effort to address the issues that are the primary subject of this paper [13]. Whilst Watson *et al* used a sensitivity of 71%, which appears consistent with the previously mentioned literature, their assumed specificity of 95% was based on laboratory test data. Even if Watson’s assumption was correct, the results of the present study remain valid. Nonetheless, it appears that the confusion matrix of table 2 from Ai *et al* ‘s study [3] is still the best estimate of the characteristics of RT-PCR testing for SARS-CoV-2 in the field. Moreover, it will still be shown in section 4.1 that Watson’s assumption, and the USA specificity reverse-engineered in section 3.3, still lie below what is needed for the use cases of the RT-PCR test.

### 2.1. False Negatives

The data from Ai revealed that more than 1*/*3 of infected people would be expected to return a negative result and return to their usual routine. Even if this person develops symptoms later, their negative test result provides false security that may affect their behaviour towards the risk of spreading the disease. False negatives can occur when not enough virus material is present in the sample, either due to the biological response of the patient or the sampling. They could also occur due to incorrect processing of the sample. The principal danger with false negatives is that an infected patient is considered safe and potentially infects others. There is evidence of multiple false negatives that proved challenging and time consuming to diagnose [19] — repeating the test may not be a valid solution (the statistical independence of multiple tests will be addressed shortly).

It appears that there is some time dependence as one would expect, as reported by Kucirca [20] and shown in figure 2.1. Virus shedding is extremely low at the moment of infection, the sensitivity first passes above 50% around 4-5 days after exposure, reaching a peak at 8 days, before decreasing slowly. This goes some way to explain the challenges faced by multiple negative test results in a patient admitted for hospital care [19]. The timing of the testing with respect to the progression of the infection is therefore quite important, and is in many ways linked with complex social factors. A test subject may be motivated to seek or avoid testing at certain times; they may report erroneous timings of symptom onset due to poor memory or optimistic estimation; or they may be pressured to take a test at an inopportune moment. The worst-case scenario from figure 2.1 is someone taking a test in the early stages of infection and almost certainly receiving a negative result. They then believe themselves to be free of the disease even as symptoms develop, and do not take the necessary precautions to avoid spreading the infection to others. Even at the peak of sensitivity, one could expect false negative rate of 21%.

**Figure 1:**
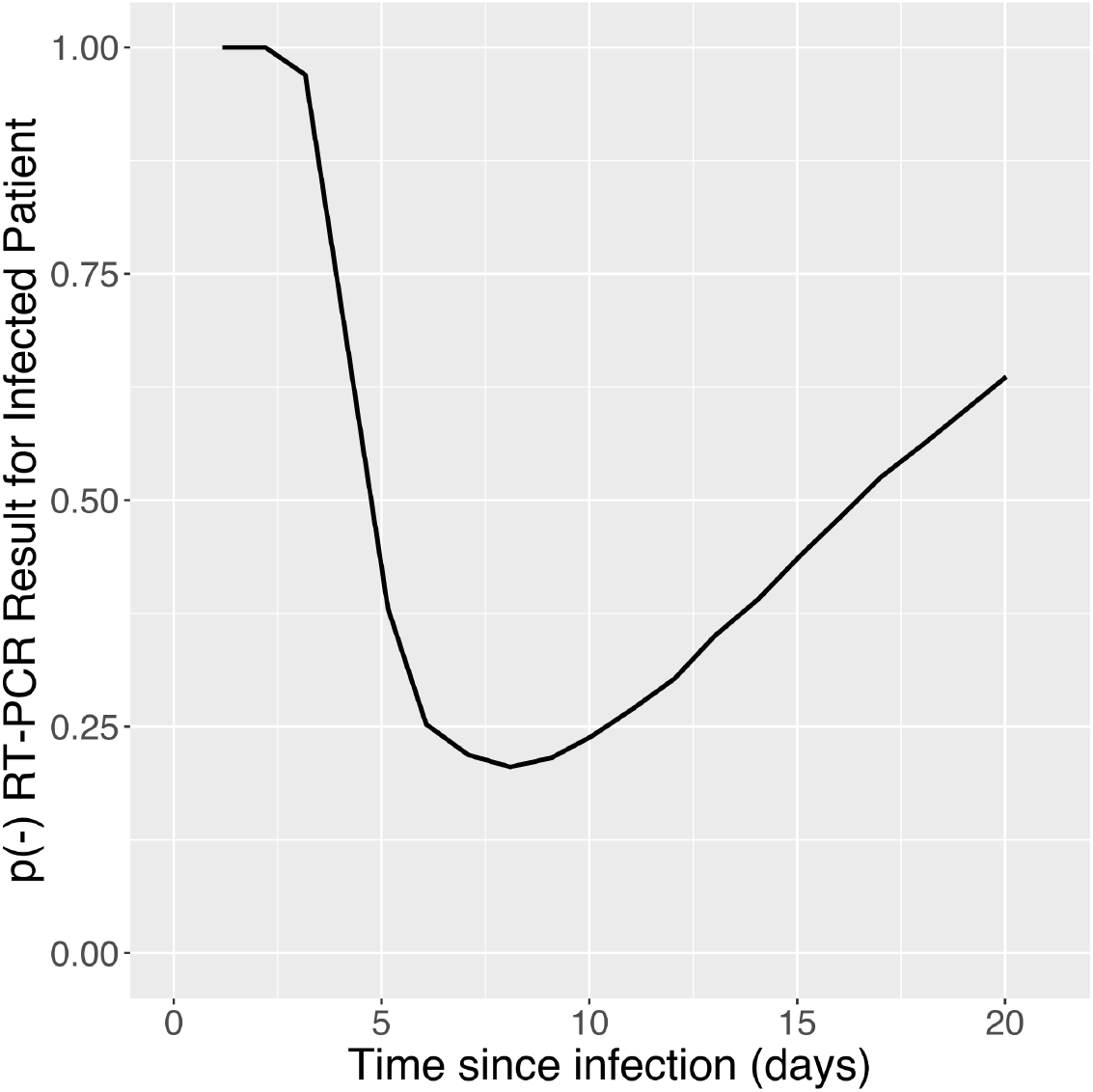
Time dependence of the probability of obtaining a negative RT-PCR result from an infected patient, after Kucirca *et al* [20].

**Figure 2:**
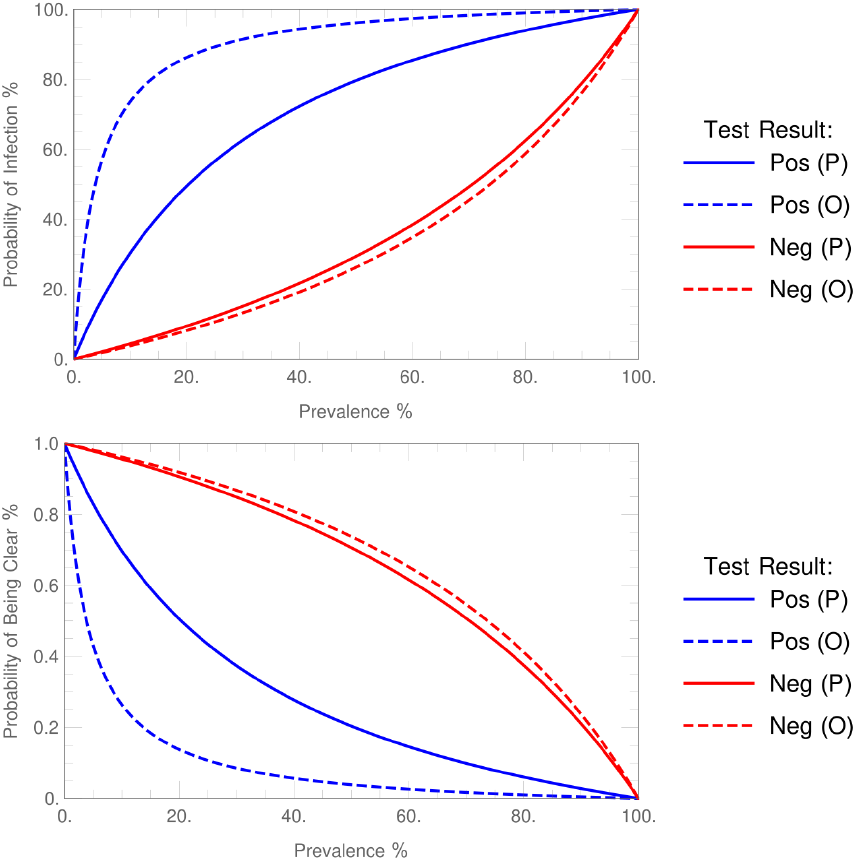
The probability of a correct RT-PCR test result, for both positive and negative test results, *vs* the prevalence of SARS-CoV-2 in the test pool per 100k population. Two curves are given for each, where (P) indicates a pessimistic 17% false positive rate, and (O) indicates optimistic 2.6% false positive rate. At the time of writing, many western countries are experiencing a prevalence of 0.6% (600 cases per 100k population in a 2 week period).

After 16 days, the sensitivity drops back below 50% again. Of course, at some point the lack of measurable virus presence transitions from a “false negative” to a status of recovered health. Many regions have assumed a 14 day quarantine period, and a 14 day reporting statistical window, which are compatible with this curve. The average of this curve is broadly consistent with Ai’s data, and so the remainder of the study will use a single false negative rate for simplicity.

### 2.2. False Positives

The “Healthy” column of table 2 shows the specificity implied by the Ai *et al* data. Almost 1*/*5 of healthy people would be incorrectly identified as infected.

False positives could occur with contamination and incorrect processing of the sample, amongst other mechanisms [21]. Large “batches” of false positives have been tied to specific test kits [22], and how they were used [23] (although, as part of that explanation it appears that there is a misunderstanding of the false negative rates). A major risk scenario is admitting a sick patient, who tests positive, into a SARS-CoV-2 ward when they actually have a different illness, a situation that was narrowly avoided in Japan recently [24]. Fortunately, a clinical assessment intervened and the patient was separated from SARS-CoV-2 patients pending further investigation.

False positives are less dangerous in wide screening settings — unlike some Kafkaesque drug testing scenarios, for example — but false positives raise anxiety and carry social and economic costs that spread into the community around those tested. There is a risk that a false positive result creates an understandable yet mistaken belief in possessing some immunity, leading some to potentially place themselves and their close contacts at increased risk of infection. False positive test results might also affect plans for vaccination: if a significant fraction of positively-tested patients have no detectable antibody level, this might be misunderstood as a loss of immunity rather than incorrect test results. The same applies for anecdotal stories of people who report having had mild SARS-CoV-2 in spring, then recovering, only to suffer a severe SARS-CoV-2 illness later in the year. Some of those cases may be false positive test results.

More recent indications of false positive rates [21] indicate possible improvements may have been made, raising sensitivity and specificity above 95%. However, on further examination of the cited references (*e.g*. [25, 26]) one finds that these are again *laboratory* studies rather than clinical studies. Mayers and Baker [25] state that in the UK, the *operational* false positive rate is unknown. Most recently, Cohen *et al* reviewed [27] the available literature and found two clinical studies reporting false positives. The first by Albendin-Iglesias *et al* [28] indicates clinical false positive rates of around 2.6% (CI 0.9-4.3%). The second by Katz *et al* [29] reports the use of multiple tests and a clinical false positive rate of 7.1% with disruption to planned medical procedures as a result. Unfortunately, in the Katz *et al* publication it does not appear that full a data breakdown of cases and test results is given, with which to estimate the confidence interval.

It will be shown in section 3.3 that the clinical specificity in the USA is generally above 91% (*i.e*. the false positive rate is below 9%).

### 2.3. Working Confusion Matrix

The working confusion matrix for this study uses the sensitivity data implied by Ai *et al* without modification. Regarding the clinical specificity, there appears to be more variation. One has a false positive rate of:

- 16.7% (CI 10–23%) from Ai *et al* [3]
- <9% from section 3.3
- 7.1% from Katz *et al* [29]
- 2.6% (CI 0.9–4.3%) from Albendin-Iglesias *et al* [28]

Henceforth, two figures will be generally given, as a range. The pessimistic is the data of Ai (*≡ P* −#), and the optimistic is the data of Albendin-Iglesias (*≡ O*−#).

### 2.4. Priors

Once one has established a confusion matrix for the test, one must then estimate the prior, or pretest, probability of being infected (from the prevalence) and some conditional probabilities of shared symptoms with other illnesses such as colds and influenza.

One therefore requires answers to the following questions:

1. What is the prevalence of SARS-CoV-2, or what is the probability of being infected by SARS-CoV-2 within a given time window (e.g. 14 days)?
2. Of those infected with SARS-CoV-2, how many have symptoms matching colds or influenza?
3. Of those infected with SARS-CoV-2, how many have symptoms that are unique indicators of SARS-CoV-2 infection?
4. What is the probability of being infected by colds or influenza within a given time window (e.g. 14 days)
5. What is the probability of suffering serious symptoms (e.g. pneumonia, CT anomalies) whilst infected with colds or influenza?

Regarding the first point (q. 1), the infection rate has been tracked by ECDC. One assumes these data are mainly positive RT-PCR test results, and at the time of writing these placed many western countries around 600 cases per 100,000 citizens in a 14 day window at the autumnal “second wave” peak in many western countries (= 0.006) [30]. This was still a low prevalence rate, far below the error rates of the test. The cumulative of the rate would be proportional to the seroprevalence as studied by Eckerle and Meyer from several hot-spots [31]. One sees at most a seroprevalence of just over 7% in Sweden, and a somewhat higher level above 10% in the most infected areas around Madrid and Geneva, after a few months of the disease spreading. In light of those figures, an average 14 day infection rate of 600 cases per 100,000 citizens seems reasonable.

Some of these other questions are partially answered by a study of passengers aboard the cruise ship “Diamond Princess” [32]. Around 54% (CI 50–57%) showed cold-like symptoms (q. 2) at the time of testing, around 10% (CI 7– 12%) required intensive care (q. 3) and 2.4% (CI 1–4%) died (note that there is an error in their paper). These numbers are about to be challenged, somewhat, in the next section.

In the absence of SARS-CoV-2, the symptoms of cough and fever together would indicate influenza, but this correctly identifies influenza around 2/3 of the time [33]. Clearly, the use of mild respiratory tract infection symptoms is not reliable in distinguishing between SARS-CoV-2, common colds and influenza.

Regarding more unusual mild symptoms, a recent study by Bénézit *et al* [34] linked positive corona tests in France with hyposmia and hypogeusia, with a sensitivity of 42% and specificity of 95%. However, both of these symptoms are not specific to SARS-CoV-2. Indeed, a study pre-SARS-CoV-2 by Henkin *et al* [35] reported around 61% of influenza patients reporting anomalous taste and smell effects. Moreover, Bénézit’s study filtered SARS-CoV-2 patients using RT-PCR results! This study should be considered inconclusive in light of the present article, but a similar study focussing on patients admitted to hospital and subject to a more rigorous assessment would be most interesting.

There are some anecdotal links reported between dysgeusia and possible SARS-CoV-2 infection, where a metallic/sour taste is experienced with the other common cold symptoms (including by this author, which resulted in this article). Lozada-Nur *et al* [36] and Aziz *et al* [37] have reviewed the literature on this topic and suggest that it may be a rather common symptom, but unfortunately these studies did not isolate dysgeusia specifically and bundled all the sensory disturbances under a common bracket. One therefore, regretfully, must ignore for now the symptoms as a distinguishing factor.

Question 4 is answered by Eccles [38], and is in the range 2-5 per year. The calculations in the present study use 4/yr as a working number. Assuming each cold/flu lasts on average a week, one can scale 4/yr to compare with 14 day infection rates of SARS-CoV-2. This 14 day cold rate (15%) is the prior that will be used for common colds and influenza.

Question 5 has been tracked by the US Centers for Disease Control and Prevention (CDC) [39] where, for example, the 2017-2018 influenza season resulted in a hospitalisation rate of 1.8% and a death rate of 0.14% out of a total of around 44.8 million cases for influenza.

As mentioned earlier, a number of studies have used RT-PCR tests as a “gold standard” reference, without referring to the matrix of confusion as given in table 2. Therein lies our problem. For example, if the entire Diamond Princess population of 3711 people were healthy, then a RT-PCR test campaign will nonetheless return approximately P-619 or O-96 positive results (all false). In reality, 712 tests were returned positive, indicating a non-zero infection rate on the ship, but the number of infected people was clearly not 712.

If one were to look at country data, for example Sweden, the European Centre for Disease Control (ECDC) reports [30] that 1000-2500 tests were performed per week per 100,000 population. Assuming the number is at the low end of that range, this is a total of 100,000 tests per week for a population of ∼ 10 million. Were the whole population healthy, one would record 16,667 false positives per week, which is P-2381 or O-371 false positives per day. This should be compared with the daily reported case rate averaged over 14 days for the same period, i.e. 4007 cases per day. Again we see that the actual infection rate is non-zero, but the false positive rate of the RT-PCR test would suggest that the real infection rate is lower than the reported cases.

## 3. Results

### 3.1. Correction of Diamond Princess Data

The pessimistic estimate of specificity is appropriate in this case, since the work was done early in the pandemic and likely used similar RT-PCR kits to those used by Ai *et al*. Using the correction equation 9 and the pessimistic specificity, we are solving the simultaneous equations:

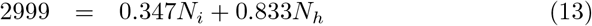

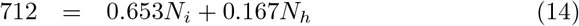

Solving yields the total number of infected patients aboard Diamond Princess to be *N*_*i*_ = 192; of which 37 patients required intensive care (≈19%, CI 14–25%) and there were 9 deaths (≈ 5%, CI 2–8%). The remaining 189 symptomatic patients were possibly suffering from a different infection spreading through the ship. The false positive rate may also explain why passengers who had been isolated in their rooms were reported to be testing positive — at the time the air ventilation systems were hypothesised to be responsible for the transmission, but for some of those patients it is likely that the false positive rate of the test is a more plausible explanation.

Tabata *et al* [40] reported that 107 people were taken to a military hospital after returning positive RT-PCR tests, and the fortunes of 104 patients were followed after 3 withheld consent. 33/104 were asymptomatic at the end of the observation period; 43/104 had mild symptoms and 28/104 had more “severe” symptoms. Of the 33 asymptomatic people, 17 had abnormal radiographical lung findings which are linked with SARS-CoV-2 diagnosis [3]. Of the 71 symptomatic patients with positive RT-PCR results, 52 (73%, CI 63–84%) had abnormal lung radiographical findings.

From these data, it appears that Tabata *et al* ‘s study has captured at least 52+17=69 of the ∼ 193 infected patients. These figures indicate that symptom-free SARS-CoV-2 may be around 17/69=25% of cases (CI 14–35%) — and conversely 75% (CI 65–86%) of patients exhibit symptoms, in answer to q. 2.

### 3.2. Sweden

Likewise for the previous subsection, equation 9 yields *N*_*i*_ = 3336 infected people per day, slightly lower than the official count of 4007. Swedish state television reported daily intensive care admissions [41] at 190 per day at the time of writing, which is 5.6% of cases. The death rate in Sweden was 19 per day, suggesting 0.6% mortality rate. These are much less intimidating figures, with a broader social demographic, in comparison to those of the cruise ship, though the Swedish figures were increasing through an autumnal “second wave” and both hospitalisation and death tend to be delayed [42], by a median of 12 days and 19 days respectively.

Taking these delays into account, one should look at the case rates over the time window of 2-4 weeks prior, at which time there were a corrected *N*_*i*_ = 1147 infections per day at the start of November 2020, implying that around 17% of patients will require intensive care, and a mortality rate of approximately 1.7%. These are at the lower end of the range of confidence of the Diamond Princess cases.

If one uses the optimistic specificity of 97.4%, the corrected infection rate increases to *N*_*i*_ = 5797 per day, higher than the official 4007 case rate because of the false negative rate. Time shifting 2-4 weeks prior, one obtains *N*_*i*_ = 4100, coincidentally similar to the official up-to-date case rate. This would imply 4.6% require intensive care, and a mortality rate of 0.5%. These seem anomalously low. There are a few possible explanations:

- The test false positive rate in Sweden was much higher than the optimistic rate (most likely explanation)
- Swedish medical care provided outlooks that are significantly superior to the those of the Diamond Princess population (unlikely)
- The virus in Sweden had evolved to a less dangerous form than experienced by those infected on Diamond Princess (unlikely)

From this, it seems logical to conclude that in Sweden the false positive rate for RT-PCR is *significantly* higher than the optimistic rate, and closer to the pessimistic values in section 2.3.

### 3.3. USA

The US CDC [11] reported 79,611,982 tests, of which 6,873,739 were positive. Applying equation 9 to these data with the pessimistic specificity indicates a negative *N*_*i*_. This can only happen if the model false positive data are too high for the USA. This is encouraging. Calculating *N*_*i*_ as a function of specificity, one sees that *N*_*i*_ first becomes positive for a specificity just above 91%, suggesting that — in the USA at least — the false positive rate is less than half of the pessimistic estimate in section 2.3, and that the approach proposed by Watson *et al* [13] to use the laboratory specificity rates of 95% are close to the operational parameters in that case.

The optimistic specificity yields a solution *N*_*i*_ = 7, 659, 736, again this is higher than the official count because it corrects for the false negative rate.

### 3.4. Bayesian Inference

#### 3.4.1. Summary of Priors

The accumulated prior probabilities from the first half of this article are summarised in table 4. Note that the entry “Cold/flu Rate” combines both the illness rate and the probability of exhibiting symptoms.

**Table 4:** Parameters used in the Bayesian analysis. “Cvd” here denotes SARS-CoV-2. *r*_*c*_ of 0.006 corresponds to 600 cases per 100,000 people in a two week period.

| Parameter | Symbol | Value |
| --- | --- | --- |
| 14d Cvd Rate | $r_c$ | 0.006 |
| 14d Cold/flu Rate | $r_f$ | 0.15 |
| Cvd Symptoms if infected | $s_c$ | 0.75 |
| Cvd hospitalisation rate | $h_c$ | 0.19 |
| Flu hospitalisation rate | $h_f$ | 0.018 |

Armed with these data, one can proceed to examine scenarios such as “If someone has a cough, and receives a negative RT-PCR test result, how probable is it that they do *not* have SARS-CoV-2 and are able to return to work?” or “If we test a person who appears healthy, and they test positive, what is the probability of infection?”

#### 3.4.2. Corrected RT-PCR Test Curves

Taking into account the base rate and marginal probability, and using the pessimistic specificity in section 2.3, the probability of a correct test result *vs* the SARS-CoV-2 prevalence is shown in figure 2. There one can see that, at a prevalence causing alarm (600 cases per 100k population), the positive RT-PCR tests almost always yield incorrect results. The negative curve, on the other hand, matches that of Woloshin *et al*, and they have a good online figure for interested readers to explore the maths with different levels of sensitivity and specificity.

These curves are “blind tests”: one tests everyone, irrespective of symptoms or other factors. In the following sections, Bayesian inference will be applied to combine sequentially the effects of reporting symptoms in combination of taking tests for some scenarios of interest.

#### 3.4.3. Mild Symptoms and Positive Test Result

The first example is a person from a social pool with 600 cases per 100k population, who has only mild symptoms and either they are requested to take a test because of employment, or they are worried. The analysis is shown in table 5. Without a test, they have a 2% probability of being infected by SARS-CoV-2, and with a positive test result this increases to an 11–42% probability of being infected, depending on whether one uses the pessimistic or optimistic false positive rate respectively. As a result, 58–89% of such people will believe they have corona without actually having the disease. Any antibody studies performed on these individuals later will be erroneous, because it is unlikely that any antibodies will be detected.

**Table 5:** Bayesian inference of a positive SARS-CoV-2 test on a person with cold/flu symptoms, assuming 600 cases per 100k population. The final, posterior probability of SARS-CoV-2 infection is 11% with the pessimistic false positive rate, and 42% with the optimistic number.

| Description | Posterior | Likelihood | Prior | Marginal |
| --- | --- | --- | --- | --- |
| Baseline / prior |  |  | 0.006 |  |
| + Cold/flu symptoms | 0.0286 | 0.754 | 0.006 | 0.16 |
| + Positive test (P) | **0.11** | 0.653 | 0.0286 | 0.181 |
| + Positive test (O) | **0.42** | 0.653 | 0.0286 | 0.0439 |

#### 3.4.4. No Symptoms and Positive Test Result

The next patient to consider is someone from a social pool with 600 cases per 100k population who has no symptoms, but they take a test either as a mass-screening project or because through a tracing system someone they have contacted was identified as being positive for SARS-CoV-2. The analysis is shown in table 6. Before testing, this person has a 0.1% probability of being infected. After a positive test, they have a 0.6% – 4% probability of being infected. This person also represents a spurious data point in any future research, since they most likely do not possess any immunity.

**Table 6:** Bayesian inference of a positive SARS-CoV-2 test on a person with no symptoms, assuming 600 cases per 100k population. The final, posterior probability of SARS-CoV-2 infection is 0.6% with the pessimistic false positive rate, and 4% with the optimistic false positive rate.

| Description | Posterior | Likelihood | Prior | Marginal |
| --- | --- | --- | --- | --- |
| Baseline / prior |  |  | 0.006 |  |
| + No symptoms | 0.00148 | 0.246 | 0.006 | 0.995 |
| + Positive test (P) | **0.00580** | 0.653 | 0.00148 | 0.167 |
| + Positive test (O) | **0.0360** | 0.653 | 0.00148 | 0.0269 |

#### 3.4.5. Severe Symptoms

This patient from a social pool with 600 cases per 100k population is admitted to hospital complaining of severe symptoms and are immediately given a test. The analysis is shown in table 7. Before testing, the patient has a 29% probability of being infected. If the test returns a positive result, they have a 62–91% probability of being infected (depending on the false positive rate), and if negative they have a 13–15% probability of being infected.

**Table 7:** Bayesian inference of a positive or negative SARS-CoV-2 test result from a person admitted to hospital with severe symptoms, assuming 600 cases per 100k population. The final, posterior probability of SARS-CoV-2 infection is 62–91% for the positive test result, and 13–15% for the negative test result, depending on whether one is pessimistic or optimistic regarding false positives, respectively.

| Description | Posterior | Likelihood | Prior | Marginal |
| --- | --- | --- | --- | --- |
| Baseline / prior |  |  | 0.006 |  |
| + Severe symptoms | 0.294 | 0.192 | 0.006 | 0.0039 |
| + Positive test (P) | **0.620** | 0.653 | 0.294 | 0.310 |
| + Positive test (O) | **0.913** | 0.653 | 0.294 | 0.211 |
| Baseline / prior |  |  | 0.006 |  |
| + Severe symptoms | 0.294 | 0.192 | 0.006 | 0.0039 |
| + Negative test (P) | **0.145** | 0.347 | 0.294 | 0.690 |
| + Negative test (O) | **0.129** | 0.347 | 0.294 | 0.789 |

#### 3.4.6. Exposed Person No Symptoms

This person was taken from an outbreak pool where 2/3 of people are infected. The analysis is shown in table 8. Before testing, the patient has a 36% probability of being infected. After a negative test result, they have a 17–19% probability of being infected, depending on the false positive rate. Almost 1/5 of the “cleared” patients will actually have the infection. On the other hand, a positive test result indicates a 69–93% probability of being infected for pessimistic and optimistic false positive rates respectively.

**Table 8:** Bayesian inference of a SARS-CoV-2 test on a person taken from a social group with high prevalence and no symptoms. The final, posterior probability of SARS-CoV-2 infection is 17–19% for a negative test result, and 67–93% for a positive test result, depending if one assumes a pessimistic or optimistic false positive rate, respectively.

| Description | Posterior | Likelihood | Prior | Marginal |
| --- | --- | --- | --- | --- |
| Baseline / prior |  |  | 0.666 |  |
| + No symptoms | 0.361 | 0.246 | 0.666 | 0.450 |
| + Negative test (P) | **0.190** | 0.347 | 0.361 | 0.658 |
| + Negative test (O) | **0.167** | 0.347 | 0.361 | 0.658 |
| Baseline / prior |  |  | 0.666 |  |
| + No symptoms | 0.361 | 0.246 | 0.666 | 0.450 |
| + Positive test (P) | **0.689** | 0.653 | 0.361 | 0.342 |
| + Positive test (O) | **0.934** | 0.653 | 0.361 | 0.252 |

#### 3.4.7. Exposed Person With Symptoms

This person with symptoms was taken from an outbreak pool where 2/3 of people are infected. The analysis is shown in table 9. Before testing, the patient has a 91% probability of being infected. After a negative test, they have a 77–80% probability of being infected. This is perhaps the most challenging scenario. This person could be “cleared” by the test under some current policy scenarios. Keeping them quarantined protects others, but 20–23% of the patients are expected to be clear of SARS-CoV-2 and holding them back puts them at risk of infection.

**Table 9:** Bayesian inference of a negative SARS-CoV-2 test on a person with symptoms taken from an infected group with high prevalence. The final, posterior probability of SARS-CoV-2 infection is 77–80% with a negative test, and 97–99.5% with a positive test, depending on a pessimistic or optimistic assumed false positive rate, respectively.

| Description | Posterior | Likelihood | Prior | Marginal |
| --- | --- | --- | --- | --- |
| Baseline / prior |  |  | 0.667 |  |
| + Symptoms | 0.905 | 0.754 | 0.667 | 0.550 |
| + Negative test (P) | **0.798** | 0.347 | 0.905 | 0.393 |
| + Negative test (O) | **0.772** | 0.347 | 0.905 | 0.407 |
| Baseline / prior |  |  | 0.667 |  |
| + Symptoms | 0.905 | 0.754 | 0.667 | 0.550 |
| + Positive test (P) | **0.974** | 0.653 | 0.905 | 0.607 |
| + Positive test (O) | **0.995** | 0.653 | 0.905 | 0.593 |

On the other hand, after a positive test, they have a 97–99.5% probability of being infected.

## 4. Discussion

It is a known fact that low prior probabilities have a significant impact on posterior probabilities, but nonetheless the worked examples should be a guide to informed decision making for likely scenarios.

At low prevalences, even if the test result is positive and one assumes that the false positive rate is at the most optimistic end of the range, whether the patient has symptoms of respiratory tract infection is the differentiating factor, taking the infection probability from 3.6% to 42%, as shown in tables 5 and 6. Nonetheless, more than half of those testing positive and having mild symptoms will still not be infected! Scientific studies using these patients can not be relied upon, unless some other expert input has been given in the diagnosis. Such a clinical diagnosis might include, for example, taking into account contact with a person who has exhibited more severe SARS-CoV-2 symptoms and had a positive test.

At the other end of the prevalence scale, one sees that in a group with 2/3 assumed infection prevalence, a negative test result with no symptoms carries just less than 20% risk of infection, whilst mild symptoms with a negative test result indicates just under 80% infection risk. Once again, it is the presence of symptoms that affects the probabilities more than the test result alone, and knowing that there is a delay of almost a week before the onset of symptoms those patients should still be quarantined. For positive test results in this pool, the presence or not of symptoms becomes irrelevant.

There are anecdotal stories of people being offered repeat tests in order to reduce the error rate for the combined results. For example, let us assume that the false negative rate is 35% and the first test is negative (ignoring prevalence and symptoms). The test is repeated and it is also negative. The assumption at this stage is that the false negative rate is 0.35 ×0.35 = 0.123. This is incorrect, because the false negative rate may be a systematic error due to the virus shedding mechanics [20], collection of sample, and its processing — the two tests are not stochastically independent. The same is true in the effort to guard against false positives: if the test kits both come from the same batch, are processed by the same people, in the same facility, using the same “black box” procedure, then they are unlikely to be stochastically independent and the errors in both tests are correlated. It is a standard procedure in science and engineering that the validation of any result be truly independent, for this reason. It would take an expert eye with experience in RT-PCR to look at the fluorescence *vs* cycle curves to guard against the false positives in this scenario, which appears to be the key to Australia’s successful testing programme (see next section).

From figure 2, one might think that as the disease spreads the positive test results will become more reliable. Whilst that is true, bear in mind that, in February 2020, the total adult critical care capacity of England was 4122 beds [43]. If one takes the ICU rate, computed for Sweden at around 17%, and from figure 2 a prevalence of 5,000 – 20,000 cases per 100,000 population (=2.7– 10.8 million cases in the UK) then ∼ 460,000 – 1.8 million ICU admissions would be needed for half of the positive test results to be accurate in a general mass testing campaign. This does then beg the question as to what kind of test characteristics one needs?

### 4.1. Alternatives and Required Test Characteristics

There are two primary use cases:

1. Reliably identifying infected people in the low prevalence population to isolate and reduce the spreading of the disease.
2. Reliably clearing non-infected people, in high prevalence settings, to allow them to escape from the high risk situation, or to return to essential work or education.

For use case 1: the null hypothesis is that the person is *not infected*, the test procedure aims to provide sufficient evidence to reject the null hypothesis and demonstrate a high probability of infection. This means that the satistical coincidence of healthy people testing positive needs to be low. The requirements for this test are shown in figure 3, where one sees that a false positive rate needs to be far below the prevalence — the intuitive result. A false positive rate of < 0.001 is needed to identify positive cases reliably, which corresponds to a specificity of > 99.9%.

**Figure 3:**
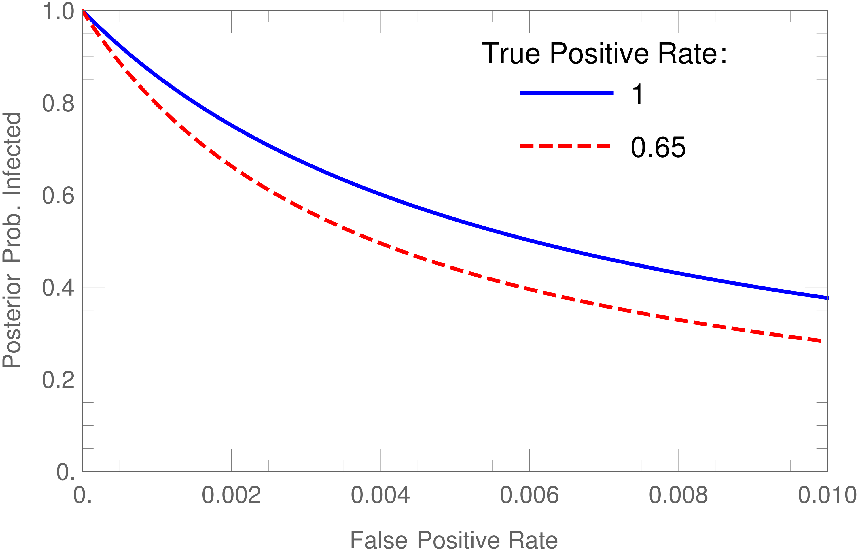
With even a relatively high prevalence of 600 cases per 100k population, these curves show that a false positive rate of < 0.001 is needed for a useful test, i.e. a specificity of > 99.9%. This result is not strongly affected by the true positive rate, as shown by the two curves indicating a perfect test or with the true positive rate of 0.65 as used in the rest of this paper.

Such figures are not inconceivable. Australia has performed a total of 9 million tests, of which a total of 1% returned positive results, which implies that under the right conditions the specificity of RT-PCR can be excellent. Indeed, informal commentary from an Australian scientist [44] explains why a black-box approach to test protocols with arbitrary thresholds will produce erroneous results, whereas an expert in RT-PCR testing would use their judgement and experience in running the apparatus. The variation in operational test characteristics in section 2.3 might be a reflection of our attempts to scale technical laboratory work beyond the hands of scientific competence, or issue performance targets and instructions to “take shortcuts”, in order to deal with an unusually high workload.

For use case 2: the null hypothesis is that the person *is infected*, and the test procedure aims to provide sufficient evidence to reject the null hypothesis and demonstrate a low probability of infection. The statistical coincidence of infected people testing negative therefore must be low. The requirements for this test are answered in figure 4. In this case, a high prevalence of 0.6 is used. One can see that a false negative rate of < 0.05 is needed to clear non-infected people at 95% confidence, matching the sensitivity of > 95%. This threshold is also intuitive. Given the time dependence of the virus shedding reported by Kucirka *et al* [20], such performance characteristics are inconceivable for RT-PCR.

**Figure 4:**
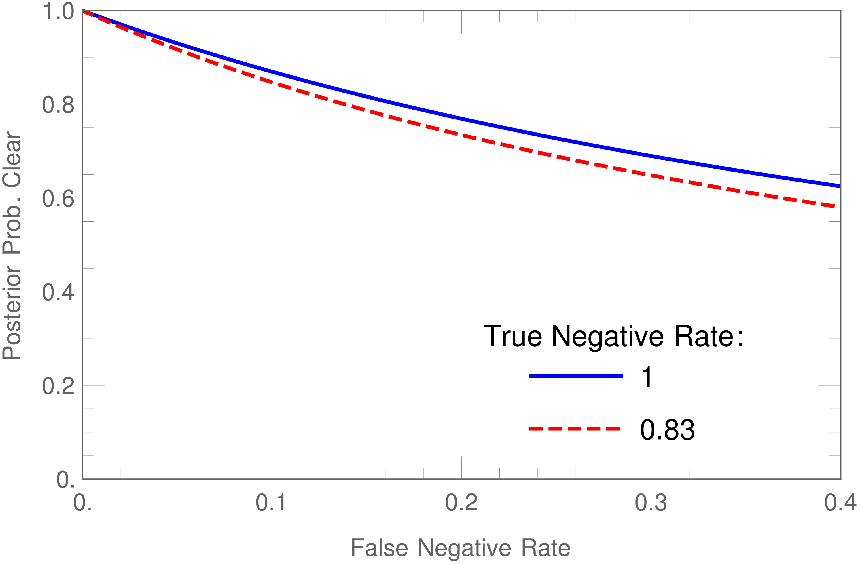
In an extremely high prevalence of 0.6 (60k cases per 100k population) such as in a hospital, jail, or some other sealed outbreak cluster, these curves show that a false negative rate of < 0.05 is needed for a useful test to rule out infection with 95% confidence, i.e. a sensitivity of > 95%. This result is not strongly affected by the specificity, as shown by the two curves indicating a perfect test (specificity = 1) or with the false positive rate of 0.17 (specificity = 0.83) as used in the rest of this paper.

Recent discussions in the literature have since turned to alternatives to RT-PCR. It is tempting, based on Ai’s study [3], to reach the uncomfortable conclusion that CT and clinical diagnosis offer a more reliable protocol than RT-PCR, a position that is refuted by Hope *et al* with good reasoning [45].

Antigen tests, whilst cheaper and faster than RT-PCR, are less sensitive and perhaps comparable in specificity when compared using RT-PCR as a gold standard [46]. This makes them useful for mass testing to estimate prevalence, but little else.

One must face the possibility that, in the short term, and based upon the mathematical nature of the problem, it is unlikely that a test exists that can reliably:

- Clear non-infected people from a pool of potentially infected people, given the low sensitivity in the early stages of infection (e.g. clearing staff and patients at medical facilities, passage at airports and regional borders)
- Identify and isolate infected people who are pre-symptomatic (e.g. finding people early before they infect others)

This viewpoint is supported by similar conclusions in the literature [18].

### 4.2. Recommendations

In future clinical studies, general at-scale RT-RCP testing alone, and tests with similar characteristics, should not be used as the ground truth SARS-CoV-2 cases. It is imperative that a more reliable diagnostic method is used, before other correlations and effects are calculated. Restricting studies to patients with hospital admissions and thorough expert diagnosis, using dedicated labs with testing experts, is likely to yield more reliable results than the non-expert, mass-testing protocols that are being used in some geographical regions.

RT-RCP tests should not be used generally to “trace” infections through individual members of the public. Whilst some countries may succeed at this (*e.g*. Australia), it depends entirely on the bandwidth of expert labs. Scaling mass testing outside expert workers [47, 48, 49] appears to be expensive and futile. Governments would do better in this way:

Step 1 Ensure the existence and support of a rigorous, dedicated, central expert group to monitor operational specificity and sensitivity of emergency use testing programmes as early as possible, on behalf of the government / region

Step 2 Use those data to correct data rates via equation 9 to *monitor the effectiveness of the strategy* to inhibit the spread of the disease in real time

Step 3 Focus the tracing efforts at targeted, critical sub-populations (e.g. medical workers, care homes, outbreak clusters) using an expert laboratories and teams dedicated to the task.

These suggestions may prove less expensive and produce more reliable results.

Negative test results (whatever the test) should not be used to “rule out” SARS-CoV-2 infection of those with symptoms or significant probability of being infected unless the test false positive rate is significantly below the prevalence. If a person exhibits symptoms of a respiratory tract infection, they should treat it with the respect it deserves and isolate themselves from society as best they can, for a duration of time based on the advice of a medical professional in their geographic location. Whether or not the infection is SARS-CoV-2, this will prevent the spread of SARS-CoV-2 and also minimise the spread of other infections that represent an enormous cost. In addition to the economic impact of the common cold, one should not forget that, globally, influenza kills millions of people each year. Such a general, isolation strategy has the added benefit of driving the circulating viruses towards lower virulence via natural selection. One can but hope that the days of sick employees demonstrating their commitment by attending work (and marketing campaigns for over-the-counter medication targeted as such) are behind us.

## 5. Conclusions

The confusion matrix of RT-PCR tests for SARS-CoV-2 has been reviewed, noting also that alternative testing kit technologies have comparable — or inferior — error rates. A simultaneous equation correction procedure for estimating the true infection rates was demonstrated for two examples: the “Diamond Princess” cruise ship and the country of Sweden in Autumn 2020, providing corrected estimates for hospitalisation and mortality rates.

Discrete Bayesian inference was then demonstrated for a few likely scenarios.

It has been demonstrated that RT-PCR testing is not reliable for three important use cases:

- RT-PCR alone can not reliably identify infected patients in a low prevalence social situation.
- RT-PCR alone can not reliably clear patients as being non-infected, if they have symptoms and come from a high prevalence social situation.
- RT-PCR alone can not reliably filter patients for subsequent medical studies such as antibody tests, symptom correlations studies, or new test candidates.

The results of this study are not entirely discouraging. Recent concern over the lifetime of SARS-CoV-2 antibodies, occasional anecdotes about repeat infection, and the need for repeated vaccination, probably need to be adjusted to take into account that many patients identified as recovered from SARS-CoV-2 who do not show measurable levels of SARS-CoV-2 antibodies are possibly associated with false positive test results in some regions (58–89% of people with mild symptoms and positive RT-PCR test results, for example). This may lead to real world antibody retention from vaccines exceeding initial expectations.

## Data Availability

Data used is all from peer-reviewed, referenced sources as stated in the article itself.

## 6. Acknowledgements

The author is extremely grateful to O. Kirstein (ESS) and Dr L. McGrath (ret) for useful scientific discussions.

